# Weighted Prevalence of Biochemically-Verified Substance Use in Healthy Adolescents Across the United States

**DOI:** 10.1101/2025.09.19.25336190

**Authors:** Natasha E. Wade, Yajuan Si, Susan F. Tapert, Janosch Linkersdörfer, Krista M. Lisdahl, Hailley R. Moore, Laila Tally, Biplabendu Das, Marilyn A. Huestis, Alexander L. Wallace, Ryan M. Sullivan, Veronica Szpak, Le Zhang, Laura Ziemer, Wesley K. Thompson

## Abstract

**Background and Aims:** Adolescent substance use is a significant public health concern, though the scope of the problem is difficult to ascertain given reliance on self-reported substance use information. Recent, large cohort studies in adolescents and young adults suggest underreporting of substance use. Analyses here aim to determine concordance between self-reported substance use and biochemical verification through hair samples and to estimate prevalence of the three most reported substances used by adolescents.

**Design:** Observational cohort longitudinal study design. Liquid chromatography and gas chromatography tandem mass spectrometry (LC or GC/MS-MS) were used to test hair samples for biochemically verified substance use, with results compared to self-reported substance use. Multi-step weighting methods estimated prevalence trends of cannabis, alcohol, and nicotine use over time, adjusting for discrepancies in sample representation due to recruitment demography, missed visits, and hair sample testing.

**Setting and Participants:** Data came from the U.S., nationwide Adolescent Brain Cognitive Development Study (n=11,868; age 9-10 at Baseline, age 15-16 at Wave 6). Participants were followed annually, with data here from 2016 to 2024.

**Measurements:** Hair samples objectively detected at least several days of substance use in a subsample of participants (n_samples_=11,865; n=6,133 unique participants). Participants self-reported past 3-month substance use. Sociodemographic, individual, and environmental-level factors were included in inverse propensity weights to estimate prevalence rates of cannabis, alcohol, and nicotine use in teens.

**Findings:** Concordance between self-report and toxicological data improved with age (for cannabis, ages 11-12=<1%; ages 15-16=45%). Weighted estimates of biochemically verified substance use indicated 7.1% (95%CI: 6.0-8.3) of 15-16 year olds engaged in biochemically detected cannabis use, 0.2% (95%CI: 0.1-0.4) used alcohol, and 4.7% (95%CI: 3.7-6.0) used nicotine.

**Conclusions:** Youth reported substance use patterns demonstrated improved biochemical verification with age. Biochemical verification reflects substantive cannabis and nicotine use by ages 15-16, supporting combining toxicological and self-report data to improve identification of substance use in youth when possible.

## Introduction

Recent scientific efforts across the globe enlist broad recruitment of large, diverse cohorts to investigate physical and mental health risk and resilience factors, across the US^1–3^, UK^4–6^, Europe^7^, the Netherlands^8^, and Switzerland^9^. In studies of youth and young adults, a common aim is to investigate predictors and consequences of substance use. As substance use represents sensitive information, self-report could be inaccurate in youth recruited into normative studies, especially those with parental involvement^9–11^.

Biochemical verification through toxicological samples can inform self-reported substance use, particularly hair analysis, which provides up to 3 months of detection (a much longer window of detection with less invasive methods than urine or blood)^12^. Initial data from several large, epidemiologically based studies suggest that many adolescents and young adults may underreport substance use. In one Zurich study, hair testing of young adults reveals substance underreporting by 30-60%^9^, with follow-up analyses indicating similar rates of under-reporting by the cohort four years later^13^. In the Adolescent Brain Cognitive Development Study (ABCD), 1,390 samples collected at ages 9-13 out of 11,868 enrolled participants indicated that hair testing could double identification of youth using substances^10^. These data highlight the utility of hair analysis to improve substance use estimate accuracy, which is key as the primary ABCD study aims focus on identifying the sequelae of substance use^14^. Further, hair testing of cannabis, nicotine, and alcohol indicates more than occasional use^15–17^, and therefore is more likely to be linked to a neurotoxic effect^18^. Due to the costs of hair toxicology, the ABCD study initially prioritized selecting samples for analyses from participants who reported risk characteristics for early substance use^11^, with a small portion of typical risk youth randomly selected for hair analyses^10^; more recent funding expanded testing all available samples at select waves.

Prioritization of participant characteristics and attrition in longitudinal studies can limit the generalizability of toxicology-based estimates. Probability sampling is the gold standard for representative prevalence estimation. Cohort studies, a crucial type of nonprobability sample, require adjustment of sample representation to match the target population^19,20^. Propensity weighting aims to statistically reduce biases (e.g., selection, retention) in order to increase accuracy of prevalence estimates. Weighting adjusts the influence of individual responses and is often recommended to better represent population norms when implemented using an appropriate external benchmark dataset^21^; accordingly, ABCD population weights are included in annual data releases^21^. While the ABCD sample was recruited through epidemiologically guided processes, it is not directly representative of the U.S., so population weights (computed in conjunction with the American Community Survey [ACS]) can help mitigate selection bias^22–24^. Yet this is not the only potential bias: attrition or fluctuating participation in longitudinal cohorts is expected, which may further impact generalizability. ABCD’s retention rate is 95%^25^, yet not all participants complete each wave of data collection. With carefully curated weights, it may be possible to better address multiple sources of potential sampling and selection bias.

Here we aim to leverage hair toxicology testing to better understand the general scope of substance use (beyond occasional or experimental use) in youth ages 9-16 through biochemical verification and propensity weighting to achieve rigorous prevalence estimates. In following a young, community-based cohort over time and using hair testing, we are better able to evaluate the extent to which generally healthy youth are using substances at more than experimental or low-level use. We first report concordance between hair testing results and self-reported substance use patterns in the three most commonly reported substances used by adolescents (i.e., alcohol, nicotine, and cannabis). We further assess the prevalence of positive test results after adjusting sample representation with the ABCD cohort through inverse propensity weighting, providing an estimate of the biochemically verified prevalence of these three most used substances in youth ages 9-16.

## Methods

Data were drawn from ABCD Data Release 6.0 (https://doi.org/10.82525/jy7n-g441), including all annual visits from Baseline (ages 9-10) through half of Wave 6 follow-ups (ages 15-16) in 11,868 youth. Hair testing was conducted on a subset of participants (n_samples_=11,865 from 6,133 unique participants; 52% of the cohort; see **Figure 1**). ABCD participants were enrolled from their local catchment centers through epidemiologically guided school recruitment from 2016-2018 at 21 sites across the United States, with four sites mainly recruiting participants from twin registries^26^. Study sites were selected, in part, to ensure recruitment of a cohort reflective of the demography of the U.S. population and covering all four census zones, with over 20% of youth ages 9-10 living within catchment regions^26^. Since one of ABCD’s primary aims is to study how substance use influences development^14^, the cohort was recruited at an age when they were expected to be substance naïve and continues to follow them as some initiate and escalate substance use^27^. Inclusion criteria at Baseline included living within the catchment area; being 9-10 years old and fluent in English; having no history of substance use disorder, severe mental illness, moderate-to-severe intellectual or developmental disability, or history of major neurological disorder including severe traumatic brain injury; birthweight >2 lbs; and no magnetic resonance imaging contraindication. Participants needed an English or Spanish speaking parent/guardian to consent and participate. Eligible participants attend data collection sessions at their local site annually and complete brief mid-year interviews remotely.

**Figure 1.** Visualization of Study Timeline, Ages, and Number of Hairs Samples Per Wave.

The present analyses include all participants with data from any visit (n=11,868). To estimate the biochemically verified three-month prevalence of substance use across the ABCD follow-up waves, we compute weighted hair testing results, where we combine (1) the ABCD population weights at Baseline^21^ with two sets of newly created weights to sequentially adjust for (2) whether a participant completed or missed a given wave and (3) whether hair samples were successfully assayed by drug test for that wave. We apply the final combined weight to positive hair results for cannabis, alcohol, and nicotine by wave of data collection, improving estimates of the prevalence of these three most used substances in youth ages 9-16.

### Measures

#### Hair Sample Collection and Toxicological Analysis

ABCD research assistants collected ∼100mg of hair closest to the root around the crown of the head from all participants who had hair styles that would not be negatively impacted by collection and were willing^11^, incorporating culturally-informed techniques^28^. Hair samples were securely stored in envelopes inside plastic sleeves and placed in locked cabinets in temperature-controlled rooms until selected for shipping, consistent with instructions from the testing laboratories. Upon receipt by the laboratory, hair was measured and trimmed to 3.9 cm, if necessary, and washed to mitigate against environmental exposure contamination, as recommended by the Society of Hair Testing^12^. The hair sample then underwent screening by immunoassays for most analytes and tested directly on mass spectrometry for cannabinoids and ethyl glucuronide (EtG). Presumptive positives were confirmed via liquid chromatography tandem mass spectrometry (LC-MS/MS) or gas chromatography tandem mass spectrometry (GC-MS/MS). Collection and testing details are in the Supplement.

A full drug panel was assessed; here we report on the most commonly self-reported substances used by adolescents: (1) cannabinoids (THCCOOH, the definitive indicator of personal ingestion^29^); (2) alcohol (EtG); and (3) nicotine (cotinine). The two testing laboratories employed by ABCD used different cutoffs to determine positive results across drug classes, so to harmonize results across labs, positive results were identified in several ways: for general concordance rates, each lab’s determination of a positive was used; for a more conservative estimate when applying prevalence weights (“high threshold”), quantitative results with the higher cutoff between the two labs were used; for a more sensitive estimate when applying prevalence weights (“low threshold”; presented within the Supplement), samples above the limit of detection (LOD) at each laboratory were identified. While the high threshold partially reduces sensitivity as described in prior reports^10^, it allows for more direct comparison across labs. LODs and cutoffs are detailed in the Supplement.

To select which hair samples to test, the ABCD Study originally relied on prior known risk factors for substance use (i.e., self-reporting substance use, family history of substance use disorder, peer use, curiosity about substance use, toxicological screening indicating substance use, externalizing symptoms, parent report of youth drinking, and older age)^11^, with an additional ∼5% of typical risk youth randomly selected for hair analyses^10^. However, in 2024, additional funding allowed for the testing of all hair samples available at Year 2, 4, and 6, as well as a larger proportion of Year 5 samples.

#### Self-Reported Substance Use

Trained research staff annually administered the Timeline Followback (TLFB) interview^30,31^ assessing use pattern, frequency, and quantity of all major substance categories in standard units including cannabis (flower, concentrates, edibles, tinctures, synthetics), alcohol, nicotine (tobacco products, electronic nicotine delivery systems or vapes, and nicotine replacement products), and other drug use. Participants are guided through a calendar, with each month between study sessions displayed on an app designed to log substance use. Participants then report their use on each given day, and are encouraged to use memory cues (e.g., significant dates like holidays or birthdays; logs from their social media accounts). While this provides means of summing lifetime use (with calendar logs from annual visit to annual visit) reported in standardized units of use,^27,32^ past-3-month substance use can also be calculated (e.g., through the ABCDscores package^33^) by limiting the calculation to only the three months leading up to the study visit. Here, we used the total sum of days of reported use of each substance from their annual visit going back three months.

#### Predictors

We fit logistic regression models to predict wave-specific participation and hair testing selection and use the inverse propensities as weighting adjustment factors. Predictors included in logistic regressions for creating the weights were selected based on substantive knowledge and their correlations to ensure no potential confounding factors were missed. Time-varying measures included age, parent- and youth-reported youth substance use and attitudes, youth mental health, and visit type (in-person, hybrid, or remote). Several time-invariant predictors collected at Baseline were also included: sex at birth, race, ethnicity, household income, highest level of parental education, family history of mental health disorders, the Childhood Opportunity Index, and state laws regarding recreational and medicinal cannabis^34^ (see the Supplement for a detailed list).

### Statistical Analyses

#### Concordance Between Self-Report and Hair Toxicology

Descriptive statistics were computed for participants’ self-report and hair toxicology results, including correlations between self-reported use days and drug analyte concentration. Hair testing is not expected to detect single use episodes of alcohol, nicotine, or cannabis,^15–17^ but typically requires at least several uses or more. Therefore, thresholds for number of days of use to expect a positive hair toxicology test were determined based on prior literature. For cannabis, a threshold of self-reporting at least two use days a month on average was used (i.e., ≥6 use days in past three months; consistent with prior findings for positive cannabinoid hair testing using detailed self-report and a full battery of toxicological testing in young adults^35^). For alcohol, other data in young adults suggests a couple drinks per week are required; however, authors of that study also note to use the most sensitive methods possible when using a community cohort and that hair can be used to detect lack of abstinence^36^, and so ≥2 days/month were used as a threshold for alcohol. Finally, for nicotine, a study of young adults indicate ≥1 day per month of nicotine use can result in detectable levels of nicotine^15^, with ≥1 days/month used in the present analyses.

Self-reported and hair testing rates of cannabis, alcohol, and nicotine use were compared, and analyses of the most reported substance within participants with tested hair samples (cannabis) assessed concordance of self-report and biochemical verification over time. Concordance rates were unweighted.

#### Multi-step Weighting

Propensity weighting is useful as a tool to reduce selection and other biases from a dataset to generate a more representative prevalence estimate. To do this, the created weight increases the influence on estimated prevalence of those who have a lower probability of being included in a study design, and reduces the influence of those who have a higher probability of being included.

Inverse propensity weighting was used to approximate the prevalence of positive hair drug tests, were biases due to cohort selection, attrition, and hair collection not present; through statistical adjustment, this process aims to better reflect generalizability to the U.S. adolescent population than reliance on raw numbers, though weighted results are still estimates. To obtain less biased estimates of substance use prevalence across time, we adjusted for sample representation to reduce selection bias through propensity weighting: Baseline weights, created by Heeringa & Berglund^21^ using recruitment data and an external benchmark; attrition/missing an annual visit (Wave Participation weights); and whether a hair sample was tested (Hair Test weights). Specifically, starting from the ABCD Study Baseline weights, we (1) fit logistic regression models to predict whether individuals participate in the follow-up wave and use the inverse propensity of participation as the weighting adjustment factor for attrition; (2) fit logistic regression models to predict whether hair samples were successfully tested by drug class, including as covariates selection criteria for testing and other relevant factors (e.g., sex, study site), and use the inverse propensity of being tested as the weighting adjustment factor of testing; and (3) multiplied the three weights, corresponding to Baseline selection, attrition, and hair testing, trimmed weights as necessary, and used these products as the final weights. Note that the final weight for cases with hair tested at Baseline is a product of the Baseline and Hair Test weights. All ABCD variables with potential relevance for selection for visit completion, hair testing, and substance use risk were considered for inclusion in models for Wave Participation and Hair Test weights. Each weight was then included to create the subsequent weights; the

Baseline weights were included as a predictor in the logistic regression for computing Wave Participation weights, and both Baseline and Wave Participation weights were included as predictors in the logistic regression for computing Hair Test weights. The weighting algorithms are described in more detail below, with specific variables included in the Supplement.

#### Baseline Weights

The procedure for computing the ABCD Study Baseline weights has been described previously^21^, with details in the Supplement. Their validity depends on whether the Baseline weights can substantially balance Baseline sample enrollment on factors related to prevalence of pertinent study outcomes. Baseline sociodemographic and recruitment information was used to generate propensity weights within the ABCD cohort to reflect national representation for 9-10 year-olds^21^. These weights, available in the ABCD data release and generated leveraging a high-quality external source (i.e., the ACS) that benchmarks the US population, can reduce the effect of selection bias and improve generalizability of ABCD analyses. Here, we assume that the Baseline weights have substantially accounted for selection factors related to both Baseline sample enrollment and this study’s outcomes.

#### Wave Participation Weights

Participation rate at each wave is: Wave 1 = 94.5%; Wave 2 = 92.5%; Wave 3 = 82.1%; Wave 4 = 82.1%; Wave 5 = 74.9%; Wave 6 = 42.6% (not all participants’ data are included in Release 6.0 from Waves 5 and 6). We created wave-specific weights using Baseline variables in logistic regressions to predict whether a participant participated in a given wave. Variables included were potential barriers to study visit participation (e.g., youth spending a lot of time in multiple homes; see Supplement). Wave-specific weights for each participant were computed by taking the inverse of the probability of wave participation estimated from the logistic regression.

#### Hair Test Weights by Substance

We assigned a value of 1 to a successfully assayed hair sample for the examined substance and 0 to those participating at this wave but without hair testing and fit a logistic regression to predict the testing propensity of the substance at each wave. Samples sent in for testing that had insufficient quantity to test were also given a 0 outcome. Predictors included those used to determine which hair samples to prioritize for testing^11^ and other variables that may influence substance use, created separately for each substance, given different rates of successful testing and prediction characteristics. Wave- and substance-specific hair testing weights for each participant were computed by taking the inverse of the probability of successful assaying estimated from the logistic regression. Hair Test weights were trimmed at the 95^th^ percentile to prevent the influence of extreme weights

#### Calculating Substance Use Prevalence Estimates

For prevalence estimates of the entire cohort over time for any positive hair results, data were restricted to participants who had hair testing results by substance (number of hair samples: n_cannabis_ = 11,607 ; n_alcohol_ = 11,373; n_nicotine_ = 11,720). We used the *survey* package in R^37^ to account for the final weights and family clustering effects in the prevalence estimation of any positive hair results by substance-specific hair testing results (cannabis [THCCOOH], alcohol, and nicotine) by wave. Confidence intervals of weighted estimates were obtained using the logit (Wald-type) method. Sensitivity analyses assessed trimming effects, where results including only the trimmed Hair Test weights were presented, given minimal difference between cannabis models (see **Tables S4, S5**). Boxplots of final weights by wave are provided in **Figure S1**.

Finally, in the Supplement, we estimated prevalence of *all* cannabis positives (i.e., THCCOOH, CBD, THC, Δ8-THC) in **Table S6** and prevalence of all THCCOOH positives but *not* accounting for Baseline weights (**Table S7**; i.e., adjusting to the cohort instead of the population), illustrating the crucial roles of weighting.

#### Missing Predictor Values

Missing predictor values were included as another level for each variable included in the prediction models. Continuous predictors were transformed into categorical variables (e.g., Externalizing T-score ≥65^38^), except for age and weights. Where the missingness category perfect predicted whether or not hair samples were tested, multiple imputation via the *mice* package were employed^39^: cannabis state-level policies (n_missing_=10; 0.01%), number of peers who drink alcohol (n_missing_=986; 1.4%) and get drunk (n_missing_=987; 1.4%), and using drugs for non-medical purposes (n_missing_=901; 1.3%), Externalizing T-score (n_missing_=902; 1.3%), parental education (n_missing_=14; 0.1%), and household income (n_missing_=1,017; 8.6%).

## Results

### Assayed Hair Sample Characteristics

11,865 hair samples were assayed from 6,133 participants across all 21 sites, with 1 to 6 samples per participant (1 sample=2,588; 2=1,925; 3=1,103; 4=466; 5 or 6=50); 66.8% of samples were assayed by US Drug Testing Laboratory (USDTL) and 33.2% by Psychemedics. Most assayed samples were from Waves 2 (n=3,394) and 4 (n=2,900; see **Table S8** for details). Insufficient hair quantity was available in 1,046 (8.8%) of samples to confirm testing of at least one drug analyte following a presumptive positive (see Supplement). For cannabinoid testing, 11,607 were successfully assayed; for alcohol, n_samples_=11,373; for nicotine, n_samples_=11,720 (see Supplement). Hair results across drug classes are available in **Table S9**. Sociodemographics of the tested samples and ABCD cohort, relative to ACS at the same age as Baseline, are presented in **Table 1** (breakdown by Wave in **Table S10**).

**Table 1.** Sample Characteristics of Tested Hair Relative to the ABCD Cohort and ACS Benchmark Distribution. **Notes:**Hair tested reflect Baseline sociodemographic characteristics of participants whose hair sample was tested, regardless of how many waves of hair samples were tested; ABCD Cohort reflects the Baseline characteristics of the full ABCD cohort; ACS reflects the American Community Survey sociodemographics used to derive the ABCD Baseline weights. Consistent with recommended reporting standards from NIDA for ABCD data, n’s < 10 are obscured.

| **Characteristic** | **Category** | **Hair Tested**<br>N=6,133 | **ABCD Cohort**<br>N=11,868 | **ACS**<br>2011-2015 |
| --- | --- | --- | --- | --- |
| **Sex** |  |  |  |  |
|  | **Male** | 52.4% | 52.2% | 51.2% |
|  | **Female** | 47.6% | 47.8% | 48.8% |
| **Race** |  |  |  |  |
|  | **White** | 63.3% | 66.9% | 52.4% |
|  | **Black** | 5.6% | 3.9% | 13.4% |
|  | **Asian** | 1.6% | 1.4% | 5.9% |
|  | **Hispanic** | 19.1% | 17.5% | 24% |
|  | **Other** | 10.3% | 10.2% | 4.2% |
| **Family Income** |  |  |  |  |
| | **<\$25K** | 10.5% | 13.8% | 21.5% |
| | **\$25K-\$49K** | 11.3% | 13.4% | 21.7% |
| | **\$50K-\$74K** | 11.1% | 12.6% | 17.0% |
| | **\$75K-\$99K** | 12.2% | 13.2% | 12.5% |
| | **\$100K-\$199K** | 31.1% | 27.9% | 20.5% |
| | **\$200K +** | 13.5% | 10.5% | 6.8% |
|  | **Other/Decline** | 9.2% | 8.5% | -- |
| **Region** |  |  |  |  |
|  | **Northeast** | 19.2% | 16.3% | 16.3% |
|  | **Midwest** | 15.1% | 14.7% | 21.6% |
|  | **South** | 29.7% | 28.1% | 38.0% |
|  | **West** | 36.0% | 40.9% | 24.1% |

### Self-Report and Hair Toxicology Comparison

Pearson correlations assessed correlations within drug class between drug analyte concentration and self-reported use days within those with hair samples that were positive for that drug class. Within those positive for cannabinoids, THCCOOH concentration correlated with past-90-day self-reported cannabis use days (r=.22, p<.001). Similarly, within those with positive hair samples, cotinine concentration correlated with self-reported nicotine use days (r=.17, p=.004), while EtG concentration did not correlate with self-reported alcohol use days (p=.72).

For cannabis products (see **Figure 2a**), 333 (2.8%, unweighted) reported use at least 2 times per month in the past 3 months. Products and amount of use varied, with some participants reporting up to 91 days of smoked cannabis (including vaped flower, smoked flower, concentrate use, and vaped oil), up to 90 days of edible use, and up to 33 days of cannabidiol use.

**Figure 2.** Confusion matrix displaying concordance between hair results and self-report, by drug class. Notes: All concordance rates are unweighted, with raw data presented. *+ Hair* indicates: positive hair sample based on each labs cutoff for confirmation; *- Hair*: negative hair sample; *+ Self*: self-reported substance use of 6 use days in the past 3 months for cannabis and alcohol, and 3 use days in the past 3 months for nicotine; *- Self*: less than the required use days self-reported. Percentages reflect the 11,865 samples tested across all waves.

Concordance between hair results and self-report for cannabinoids ranged was 93.9% overall, demonstrating some under-reporting of cannabis use.

For alcohol use (see **Figure 2b**), 83 (0.7%) participants self-reported 6+ alcohol use days in the past 3 months. Concordance between hair results and self-reported alcohol use was 98.1%; <10 participants were positive on hair testing and self-reported ≥6 alcohol use days in the past 3 months.

For use of nicotine or tobacco products (see **Figure 2c**), 382 (3.2%, unweighted) self-reported 3+ use days in the past 3 months. Participants reported up to daily nicotine use via cigarettes and e-cigarette/vaping products. Concordance between hair results and self-reported nicotine or tobacco use was 95.7%.

Concordance rates by substance and wave are included in **Supplemental Figure S2**.

Cannabis was most frequently confirmed on hair testing and most frequently reported amongst those with hair samples tested. At Baseline no participants self-reported ≥2 cannabis use days per month in the past 3 months and had hair positive for cannabinoids (see **Table 2**); by Wave 6, 45% (unweighted) of participants who were positive for cannabinoids self-reported ≥2x/month cannabis use days. Overall concordance (matching hair results and self-report) varies by year, with lower overall concordance in the years with the highest number of hair cannabinoid positive results. Similar data for alcohol and nicotine are presented in Tables S11 and S12, respectively.

**Table 2.** Concordance between Hair Cannabinoids and Self-Report by Wave (n_samples_=11,865) Notes: All concordance rates are unweighted. % Self-Report is the percentage of participants who self-reported cannabis use ≥2x/month by wave; % + Hair Cannabinoids is the percentage of samples tested that were positive for cannabinoids on confirmation testing; % Concordant/All Samples represent percentage of youth whose hair results matched their self-report (either positive hair results and self-reported, or negative hair results and denied use); % Positive + Concordant/Positive Cannabinoids shows that same percentage, but is restricted to those who are concordant who tested positive for cannabinoids. The lab-determined positive hair testing identification was used here. Consistent with recommended reporting standards from NIDA for ABCD data, n’s < 10 are obscured.

|  | **% Self-**<br>**Reported**<br>**Cannabis** | **% + Hair**<br>**Cannabinoids** | **% Concordance** | **% Positive +**<br>**Concordant/**<br>**Cannabinoids** |
| --- | --- | --- | --- | --- |
| Baseline | 0% | 2.5% | 97.5% | 0% |
| Wave 1 | <1% | 3.6% | 96.5% | 4.0% |
| Wave 2 | <1% | 2.8% | 97.2% | <1% |
| Wave 3 | <1% | 10.4% | 89.3% | 0% |
| Wave 4 | 1.9% | 8% | 93.3% | 19.7% |
| Wave 5 | 5.4% | 11.4% | 91.2% | 35% |
| Wave 6 | 8.1% | 11.4% | 90.9% | 45.4% |

### Weighted Hair Toxicology Estimates

Applying statistical weights (inverse propensity weights) to hair testing results provides a less biased estimate of substance use prevalence. As can be seen in **Figure 3** (full data in **Table S13**), there is an increasing prevalence rate with age in “High Threshold” weighting (more strict threshold for determining positive results; “Low Threshold” results are displayed in **Figure S2**). Weighting largely attenuates estimates, particularly in later waves when participants are older. Variation in confidence intervals likely reflects sample size differences (smaller confidence intervals are observed with larger numbers of tested hair samples) and methodological testing differences, such as the methods used within and between testing labs and the cutoff used to establish positives.

**Figure 3.** Weighted Prevalence Estimates by Substance and Wave. Notes: Shaded regions represent 95% confidence interval. High threshold refers to a higher cutoff to determine a positive hair test and a more conservative estimate.

Relative to large, important epidemiological studies, weighted prevalence varies by substance and by self-reported use frequency, though there are notable distinctions in definitions of substance use by drug class and the fact that <u>hair detects past-three month use</u>, whereas past-month is a common timeframe reported in most epidemiological studies. Another concern is the level of use required to detect substances in hair, relative to self-report in other studies; for example, Monitoring the Future (MTF^40^), alcohol used is defined as “more than a few sips” or in National Survey on Drug Use and Health (NSDUH) as more than a “sip or two”. Alcohol detection in hair requires more heavy use, which is likely still different than reports of getting drunk (queried by MTF) or binge drinking on 5 or more days (queried by NSDUH^41^). Even so, it is interesting to consider prevalence rates with these cautions. Specifically, the wave of ABCD data that most closely corresponds to 8^th^ grade (Wave 4), 6.4% had <u>hair toxicology confirmed</u> cannabis use, 0.2% with alcohol use, and 1.4% with nicotine use. In MTF, 8^th^ graders reported a <u>past-month rate</u> of 4.3% for cannabis, 4.9% for alcohol use (7.1% for 1-2 occasions; 4.7% for 3- 5 occasions; 2.6% for 6-9 occasions), 1.5% for being drunk, and 4.9% for nicotine. For NSDUH data, 6.9% of 12-17 year-olds reported any alcohol use in the past month, 3.9% binge drank, and 0.5% reported binge drinking on 5 or more days in the <u>past month</u>. For nicotine, in the National Youth Tobacco Survey, around 1.5% of middle schoolers report using e-cigarette products <u>≥6</u> <u>days/month</u>^42^.

## Discussion

Based on current raw and weighted prevalence estimates of biochemically verified substance use, substantive past-3-month substance use is present in the ABCD cohort, and for cannabis perhaps indicating more frequent use than is captured within large, epidemiological studies that do not have biochemical confirmation. Consistent with prior reports^9,10^, our data suggest some youth under-report substance use. However, concordance improves with age: while at Wave 2 (ages ∼11-12), <1% of participants were positive for hair cannabinoids and also self-reported cannabis use at a level which could be detectable on hair testing, by Wave 6 (ages ∼15-16) that rate increased to nearly half (45%) of participants. As expected, after applying statistical weights to reduce bias from study design characteristics, hair toxicology results indicate increased substance use with age, with estimated toxicologically-detected prevalence of cannabis use in 7.1%, alcohol use in 0.3%, and nicotine use in 4.7% of 15-16 year-olds. As hair testing for these substances typically requires several use episodes or more^15–17^, these data suggest more than just occasional use.

Determining the consequences of adolescent substance use requires accurate categorization of substance use onset. While studies recruiting specifically for substance use appear to show less underreporting (e.g., Wade et al.^35^), community-based studies of youth (including ABCD) appear to have misreporting^9–11,13,43^. Results indicate that, when comparing self-report to unweighted hair testing results, concordance increases with adolescent age, perhaps due to increased comfort in disclosure with age, increased knowledge of one’s substance use with age, or increased trust with study staff^44–46^. As prior results in a Zurich cohort found consistent rates of underreporting at both ages 20 and 24^13^, it may be that age benefits concordance rates up until young adulthood, wherein accuracy in reporting may plateau. At the same time, this prior work did not identify THCCOOH, nor detect alcohol or nicotine, which may limit comparisons with the present findings. In studies with capacity (e.g., financial, staff, storage), then, use of both self-report and objective, biochemical verification through toxicological data are recommended to provide the most accurate estimate of adolescent substance use^47^, including continued monitoring within the ABCD cohort.

Inverse propensity weights were used here to reduce biases in data collection from cohort recruitment, study session attendance, and hair testing selection, in order to estimate prevalence were these biases not present. Results here are helpful for establishing that non-randomized selection of biochemical verification for substance use is informative for research, though not directly generalizable. Without weighting, unweighted rates underestimate positives in early years (Waves 0-3) and may overestimate prevalence in later years (Waves 4-6). These overestimates in latter years, in particular, may reflect biases in the data (e.g., in who attends session in person, has a hairstyle which would not be disrupted by sample collection, and is willing to give a sample) which contribute to uncorrected prevalence rates which are not directly reflective of the population. Considering primary (e.g., Figure 3) and follow-up (Table S7 for assessing within-cohort generalizability) weighting models, unweighted hair data are not representative of the broader population or the full ABCD cohort, respectively. On balance, although weighting improves the comparability of the analytic samples to the baseline ABCD cohort and, through the Baseline weights, to selected population sociodemographic benchmarks, these estimates should not be interpreted as definitive population prevalence estimates. The generalizability of the weighted estimates depends on the validity of the assumptions underlying each weighting step. The Wave Participation and Hair Test weights adjust for observed differences related to wave participation and hair test selection, while the Baseline weights adjust the ABCD cohort to better match population sociodemographic characteristics using external benchmarks from the ACS. However, unmeasured factors associated with recruitment, study participation, hair test selection, or substance use may remain unaccounted for. Mechanistic studies of specific substances or analytes (e.g., THCCOOH, Δ8-THC) can reliably use hair data to confirm use, while other analyses may choose to incorporate hair data as one data point within a range of measures to identify substance use.

Weighted results for hair cannabinoids map well to other large epidemiological studies, while alcohol and nicotine in hair may be less readily detected (i.e., require more frequent and heavy use) and diminish comparisons between biochemically verified results and self-report. Notably, hair captures a larger window of use (∼3 months) than is captured in some questions from other large projects, and only detects more frequent use across substances which reduces identification of low level use, intermittent or occasional use, or subclinical levels of use^15–17,35^. Alcohol particularly requires substantial use for detection on hair testing^48,49^, explaining the low detection seen here and the discrepancies when compared with other large epidemiological studies. Therefore, results suggest broad consistency with epidemiological studies, but also that the low level of use in some self-reported and epidemiological studies (e.g., ≥1 use) may underestimate the severity and frequency of cannabis use, in particular, across adolescence.

Some factors may increase noise within these estimates. For instance, the low number of samples tested on some years, use of different labs, or within lab method changes (e.g., using GC-MS/MS for a brief period for EtG) increase noise, demonstrated through increased confidence intervals in weighted estimates (e.g., at Wave 3). Yet the totality of the pattern indicate hair toxicology data are a robust means of enhancing identification of adolescent substance use. Applying weights in this way was previously shown to reveal differential impact on neurocognition and functional imaging^23^, and may be used similarly with hair data.

Strengths of the present analyses are use of a benchmark dataset (ACS) for creating Baseline weighting^21^, broad consideration of factors potentially influencing hair testing results, focusing on metabolites (i.e., THCCOOH, EtG, and cotinine) to confirm personal ingestion of each substance, and thorough measurement of self-reported and biochemically verified substance use. Limitations are also noted. A key limitation is that the Wave Participation and Hair Test weights rely on assumptions about the measured variables that predict study participation and hair test selection. Although these weights are intended to reduce bias, there may be additional unmeasured factors influencing wave attendance, selection for hair testing, or substance use outcomes that were not captured in the weighting models. Similarly, the ABCD Baseline weights primarily align the sample with population sociodemographic benchmarks from the ACS; they may not fully account for other characteristics related to recruitment into the ABCD Study or to substance use risk. Accordingly, the weighted estimates should be interpreted as improving representativeness relative to the analytic sample, rather than as providing definitive estimates of population prevalence. Beyond this, hair samples cannot be collected from all participants, and methods are key for comfortable collection across participants^28^. Weighting results relied solely on hair drug testing results; incorporating self-report would increase prevalence estimates, as hair toxicology does not identify infrequent or occasional use. While hair color information was collected from some participants at some waves, it was at times self-reported and at times identified by staff, preventing inclusion in weights. Basic drugs are deposited in higher concentrations in hair than acidic or neutral drugs due to strong melanin binding, although cannabinoids do not^50,51^. Though there are concerns about cosmetic treatments to hair damaging detection of drug analytes, evidence suggests similar EtG, THC, and other drug analyte concentrations were found in cosmetically treated versus untreated hair^52^. Two different laboratories analyzed hair samples, with one lab changing EtG methods for a brief period of time. Considering different cutoffs, LOD, and level of quantitation (LOQ) can help inform interpretation of these results across laboratories. Other methodological differences across studies are noted. Some European studies collect smaller volumes of hair and test directly by tandem mass spectrometry rather than screening first, may not always test for metabolites (e.g., THC and CBN instead), and test to the lower level of quantitation^9,13,53,54^. Self-report questionnaires vary across all studies, including within each U.S.-based epidemiological study^40–42^ and in European studies (e.g., using a checklist to mark drugs used then reporting general frequency)^9^.

In conclusion, biochemical verification of substance use data from a diverse, large longitudinal study indicates substantive past-3-month use of cannabis (7.1%) and nicotine (4.7%) by age 15-16. This is consistent with epidemiological self-report data, and provides novel insight into higher intake levels of use in U.S. based youth. Combining toxicological data with self-report provides clear indications of more accurate self-report as youth age. Results robustly support use of biochemical verification of substance use, when available, to ensure accurate determination of substance use consequences in youth.

## Data Availability

Data is available to approved researchers via the NBDC portal

https://www.nbdc-datahub.org/

## Acknowledgements

Authors would like to thank the families who participate in the ABCD Study and the staff who collect the data. N.E. Wade was supported by National Institute on Drug Abuse (DA050779, PI: Wade; DA054980, MPI: Doran/Jacobus). This work was also supported by T32 AA013525 (PI: Riley/Spadoni to Wallace, Sullivan, and Szpak), DA062011 (PI: Wallace), and DA064409 (PI: Sullivan). Data used in the preparation of this article were obtained from the <u>Adolescent Brain Cognitive Development™ (ABCD) Study</u>, held in the <u>NIH</u> <u>Brain Development Cohorts Data Sharing Platform</u>. This is a multisite, longitudinal study designed to recruit more than 10,000 children aged 9–10 and follow them over 10 years into early adulthood. The ABCD Study® is supported by the **National Institutes of Health** and additional federal partners under award numbers: U01DA041048, U01DA050989, U01DA051016, U01DA041022, U01DA051018, U01DA051037, U01DA050987, U01DA041174, U01DA041106, U01DA041117, U01DA041028, U01DA041134, U01DA050988, U01DA051039, U01DA041156, U01DA041025, U01DA041120, U01DA051038, U01DA041148, U01DA041093, U01DA041089, U24DA041123, U24DA041147.

A full list of supporters is available at <u>Federal Partners – ABCD Study</u>. ABCD Consortium investigators designed and implemented the study and/or provided data but did not necessarily participate in the analysis or writing of this report. This manuscript reflects the views of the authors and may not reflect the opinions or views of the NIH or ABCD Consortium investigators. The ABCD data repository grows and changes over time. The ABCD data used in this report came from https://doi.org/10.82525/jy7n-g441. Additional support for this work was made possible from NIEHS R01-ES032295 and R01-ES031074.

