## Supplement for "Weighted Prevalence of Biochemically-Verified Substance Use in Healthy Adolescents Across the United States"

**SUPPLEMENTAL MATERIALS**

**Lab differences and cutoffs for determining positive hair samples**	p. 2

Table S1. LOQ/LOD by Lab

**Weighting Process and Variables**	p. 5

Table S2. Variables included in weighting factors

Table S3. Weighting with overlapping factors in each weight

Tables S4, S5. Sensitivity analyses of weight trimming

Figure S1. Boxplots of Weight Values by Wave

**Cannabinoid Positive Hair Tests**	p. 12

Table S6. Weighted Cannabinoid Prevalence Rates

**Weighted Prevalence of Wave Participation*Hair Test without Baseline**	p. 13

Table S7. THCCOOH Estimation Without Baseline Weights

**Number of samples tested and insufficient samples**	p. 14

Table S8. Number of samples tested by laboratory

**All Drug Class Positives**	p. 15

Table S9.

**Sociodemographics by Wave of Hair Tested**	p. 16

Table S10.

**Confusion Matrices of Self-Report and Toxicology by Wave and Substance**	p. 17

**Concordance Tables by Wave**	p. 18

Table S11. Alcohol

Table S12. Nicotine

**Full Weighted Prevalence Results by Substance and Wave**	p. 20

Table S13. Full Weighted Prevalence Results by Substance and Wave

Figure S2. Low Threshold Weighted Prevalence Estimates

**Lab differences and cutoffs for determining positive hair samples**

**Hair Sample Collection and Toxicological Analysis.** ABCD research assistants collected ~100mg of 3.9 cm hair closest to the root around the crown of the head from all participants who were willing and had hair styles which would not be negatively impacted by collection. Staff were trained in culturally sensitive collection methods^36^. Hair samples were securely stored in envelopes inside plastic sleeves and placed in locked cabinets until selected for shipping. The likelihood of hair testing varied over time due to changes in funding and selection criteria (e.g., the inclusion of randomly selected “low risk” participants after Wave 1 follow-up).

Between study launch and 2023, samples were processed by Psychemedics (Culver City, CA), and from 2024 onwards by United States Drug Testing Laboratories (USDTL; Des Plaines, IL). Techniques were similar across labs. Upon receipt by the laboratory, hair was measured and trimmed to 3.9 cm, if necessary. The hair sample then underwent enzymatic (Psychemedics) or alkaline (USDTL) digestion for screening by immunoassays (for most analytes) or tested directly on mass spectrometry (for cannabinoids and ethyl glucuronide [EtG]). Hair was washed to mitigate against contamination from environmental exposure, as recommended by the Society of Hair Testing ^22^. Psychemedics confirmed presumptive positive immunoassay screens via LC-MS/MS or GC-MS/MS analysis with positives set to the limit of detection (LOD)/limit of quantification (LOQ). USDTL set the LOD for ELISA screening using negative controls established in each batch of tested samples. This established a highly sensitive threshold to rule out negative hair samples, with any presumptive positive samples sent for GC/MS-MS or LC/MS-MS confirmation testing. LOD and LOQ for screening and confirmation, by drug analyte, can be seen in **Table S1**.

Labs largely used different cutoffs to determine positive results across drug classes. To better harmonize results across labs, positive results were identified in two ways: for a more conservative estimate (referred to as “high threshold” below), quantitative results with a higher cutoff were used; for a more sensitive estimate (referred to as “low threshold” below), samples above the LOD at each lab were identified. While the high threshold partially reduces sensitivity as described in prior reports ^10^, it allows for more direct comparison across labs.

Tested drug classes included alcohol, amphetamines, benzodiazepines, cannabinoids, cocaine, fentanyl, opioids, nicotine, and phencyclidine. For drug classes that can also be used as prescription medications (amphetamines, benzodiazepines, opioids, and fentanyl), participant and parent reports were used to remove positive cases that are explained by prescription use. While rates of detection across drug classes are reported within the Supplement, the Results section presents hair testing results only for cannabis, alcohol, and nicotine.

Because two different laboratories assayed collected hair samples, each used their own methodologies which, while largely overlapping, sometimes resulted in different determinations of hair positives. For Psychemedics, the cutoff was set to the LOQ which was also equivalent to the LOD. For USDTL, separate LOD, LOQ, and cutoffs were determined. Given these differences, presented positives within the manuscript were calculated in two ways, with both results presented. First, using the quantified concentration, the more conservative cutoff set by USDTL was used for all samples, regardless of testing lab. Second, a more sensitive cutoff was set to the lab LOQ of each analyte.

**Table S1.** LOQ/LOD by Lab

| **Psychemedics** | |  |  |  | **USDTL** | |  |  |  |
| --- | --- | --- | --- | --- | --- | --- | --- | --- | --- |
| **Drug Class** | **Analyte** | **LOD/LOQ** | **Cutoff** |  | **Drug Class** | **Analyte** | **LOD** | **LOQ** | **Cutoff** |
| **Nicotine** | | |  |  | **Nicotine** | |  |  |  |
|  | Nicotine | -- | -- |  |  | Nicotine | 20 | 40 | 100 |
|  | Cotinine | 50 | 50 |  |  | Cotinine | 20 | 40 | 100 |
| **Parent Cannabinoids** | | |  |  | **Parent Cannabinoids** | | |  |  |
|  | THC | 5 | 5 |  |  | THC | 40 | 40 | 40 |
|  | CBN | 5 | 5 |  |  | CBN | -- | -- | -- |
|  | CBD | 5 | 5 |  |  | CBD | 40 | 40 | 40 |
|  | THCV | 5 | 5 |  |  | THCV | -- | -- | -- |
|  | Delta-8-THC | -- | -- |  |  | Delta-8-THC | 40 | 40 | 40 |
|  | Delta-10-THC | -- | -- |  |  | Delta-10-THC | 40 | 40 | 40 |
| **THCCOOH** | | |  |  | **THCCOOH** | |  |  |  |
|  | THCCOOH | 0.02 | 0.02 |  |  | THCCOOH | 0.01 | 0.02 | 0.05 |
| **Alcohol** | | |  |  | **Alcohol** | |  |  |  |
|  | Ethyl Glucuronide | 1 | 1 |  |  | Ethyl Glucuronide | 4 | 8 | 20 |
| **Cocaine** | |  |  |  | **Cocaine** | |  |  |  |
|  | Cocaine | 25 | 25 |  |  | Cocaine | 20 | 40 | 100 |
|  | Benzoylecgonine | 25 | 25 |  |  | Benzoylecgonine | 10 | 20 | 50 |
|  | Orthohydroxycocaine | .4 | .4 |  |  | Orthohydroxycocaine | .05 | .8 | 2 |
|  | Parahydroxycocaine | .4 | .4 |  |  | Parahydroxycocaine | .012 | .8 | 2 |
| **Amphetamines** | |  |  |  | **Amphetamines** | |  |  |  |
|  | Amphetamine | 25 | 25 |  |  | Amphetamine | 20 | 40 | 100 |
|  | Methamphetamine | 25 | 25 |  |  | Methamphetamine | 20 | 40 | 100 |
|  | MDA | 25 | 25 |  |  | MDA | 20 | 40 | 100 |
|  | MDMA | 25 | 25 |  |  | MDMA | 20 | 40 | 100 |
|  | MDEA | 25 | 25 |  |  | MDEA | 20 | 40 | 100 |
| **Opiates** | |  |  |  | **Opiates** | |  |  |  |
|  | Codeine | 50 | 50 |  |  | Codeine | 20 | 40 | 100 |
|  | Morphine | 25 | 25 |  |  | Morphine | 20 | 40 | 100 |
|  | 6-MAM | 25 | 25 |  |  | 6-MAM | 20 | 40 | 100 |
|  | Oxycodone | 50 | 50 |  |  | Oxycodone | 20 | 40 | 100 |
|  | Oxymorphone | 50 | 50 |  |  | Oxymorphone | 20 | 40 | 100 |
|  | Hydrocodone | 50 | 50 |  |  | Hydrocodone | 20 | 40 | 100 |
|  | Hydromorphone | 50 | 50 |  |  | Hydromorphone | 20 | 40 | 100 |
| **PCP** |  |  |  |  | **PCP** |  |  |  |  |
|  | Phencyclidine | 100 | 100 |  |  | Phencyclidine | 20 | 40 | 100 |
| **Fentanyl** | |  |  |  | **Fentanyl** | |  |  |  |
|  | Fentanyl | 1 | 1 |  |  | Fentanyl | 2 | 4 | 10 |
|  | Norfentanyl | 1 | 1 |  |  | Norfentanyl | 2 | 4 | 10 |
|  | Acetyl Fentanyl | 1 | 1 |  |  | Acetyl Fentanyl | 2 | 4 | 10 |
|  | Acetyl Norfentanyl | 1 | 1 |  |  | Acetyl Norfentanyl | 2 | 4 | 10 |

Notes: Cutoffs, LOQ, LOD are in pg/mg

**Weighting Process and Variables**

**Baseline Weight Summary.** For the ABCD baseline weights ^15^, eight key variables (demographic, socioeconomic) which are available in both ABCD and the ACS were used: age at baseline, sex, race/ethnicity, household family income, parental marital status, parents employment status, and census region. Missing data in either the ABCD or ACS datasets were imputed. A logistic regression model was run to calculate the probability of a case being from an ABCD participant based on the catenated ABCD and ACS data. ABCD participants were more likely to have married parents and higher household income than ACS participants. Weights were computed as the inverse of the estimated probability of being an ABCD participant given that person’s socio-demographics. Weights were then trimmed at the 2^nd^ and 98^th^ quantiles to reduce the impact of extreme weights. The trimmed weights were then raked to reflect the exact ACS population distributions of age, sex, and race/ethnicity.

**Weighting Process**. When creating weights, data was first stratified by Wave. Any variables which were null (e.g., no self-reported cocaine use at Baseline) were removed from weighting models. Any variables which exhibited singularity (i.e., coefficient >|2.5|) in weighting models were examined using crosstabs and removed. Logistic models were re-run to again examine coefficients and see if the revised model after variable removal then introduced new variables with singularity, following the same process in an iterative fashion (crosstab review and removal as appropriate).

The following variables were included for creation of the weighting variable:

**Table S2**: Variables included in weighting factors

| **Variable** | **Time Variant** | **Baseline Weight** | **Wave Weight** | **Hair Weight** |
| --- | --- | --- | --- | --- |
| Age at Baseline | No | X | X |  |
| Age at Visit | Yes |  |  | X |
| Sex | Yes | X |  |  |
| Race | No | X |  |  |
| Ethnicity | No | X |  |  |
| Household income | No | X |  |  |
| Parent marital status | No | X |  |  |
| Parent employment status | No | X |  |  |
| Census region | No | X |  |  |
| Highest level of parental education |  |  | X |  |
| Surveys in Spanish for parent | Yes |  | X |  |
| Youth spends a lot of time in another home | Yes |  | X |  |
| Externalizing symptoms (>65 on CBCL) | Yes |  | X |  |
| Childhood Opportunity Index (low to high) | No |  | X |  |
| Family unable to afford food in past 12 months | Yes |  | X |  |
| Family history: AUD | No |  | X |  |
| Family history: SUD | No |  | X |  |
| Family history: Depression | No |  | X |  |
| Family history: Hallucinations | No |  | X |  |
| Family history: Hospitalization due to mental health | No |  | X |  |
| Family history: Mania | No |  | X |  |
| Family history: Trouble job/law | No |  | X |  |
| Family history: “nerve” issues | No |  | X |  |
| Baseline Weight | No |  | X | X |
| ABCD Study Site | Yes |  | X | X |
| Wave Participant Weight | Yes |  |  | X |
| Self-reported Alcohol Sipping (<standard drink) | Yes |  |  | X |
| Self-reported Alcohol use | Yes |  |  | X |
| Self-reported Cathinone | Yes |  |  |  |
| Self-reported recreational CBD | Yes |  |  | X |
| Self-reported cocaine | Yes |  |  | X |
| Self-reported DXM | Yes |  |  | X |
| Self-reported inhalants | Yes |  |  | X |
| Self-reported ketamine | Yes |  |  | X |
| Self-reported MDMA | Yes |  |  | X |
| Self-reported methamphetamine | Yes |  |  | X |
| Self-reported cannabis blunt | Yes |  |  | X |
| Self-reported smoked cannabis concentrate | Yes |  |  | X |
| Self-reported vaped cannabis concentrate | Yes |  |  | X |
| Self-reported cannabis and alcohol drink | Yes |  |  | X |
| Self-reported cannabis edible | Yes |  |  | X |
| Self-reported cannabis puff/taste (<1 standard use) | Yes |  |  | X |
| Self-reported cannabis smoke | Yes |  |  | X |
| Self-reported synthetic cannabis | Yes |  |  | X |
| Self-reported cannabis tincture | Yes |  |  | X |
| Self-reported vaped cannabis flower | Yes |  |  | X |
| Self-reported nicotine chew | Yes |  |  | X |
| Self-reported cigarette use | Yes |  |  | X |
| Self-reported cigar | Yes |  |  | X |
| Self-reported hookah | Yes |  |  | X |
| Self-reported nicotine pipe | Yes |  |  | X |
| Self-reported nicotine puff (<1 standard use) | Yes |  |  | X |
| Self-reported nicotine replacement | Yes |  |  | X |
| Self-reported vaped nicotine | Yes |  |  | X |
| Self-reported opiate use (e.g., heroin) | Yes |  |  | X |
| Self-reported other drug use | Yes |  |  | X |
| Self-reported prescription opiate | Yes |  |  | X |
| Self-reported prescription sedative | Yes |  |  | X |
| Self-reported prescription stimulant | Yes |  |  | X |
| Self-reported salvia | Yes |  |  | X |
| Self-reported mushroom | Yes |  |  | X |
| Self-reported vaped flavor | Yes |  |  | X |
| Positive urine toxicology | Yes |  |  | X |
| Positive oral toxicology | Yes |  |  | X |
| If one of your best friends were to offer you marijuana, would you try it? | Yes |  |  | X |
| If one of your best friends were to offer you alcohol, would you try it? | Yes |  |  | X |
| CBCL: Drinks alcohol without parents' approval | Yes |  |  | X |
| CBCL: Smokes, chews, or sniffs tobacco | Yes |  |  | X |
| CBCL: Uses drugs for non medical purposes | Yes |  |  | X |
| Medicinal CBD use | Yes |  |  | X |
| Breathalyzer | Yes |  |  | X |
| Marijuana state law during the same year as the assessment | No |  |  | X |
| The level of offense for possessing one ounce of MJ in a state is classified as a civil offense/infraction/petty offense or not an offense | No |  |  | X |
| How many of your friends: Use marijuana | Yes |  |  | X |
| How many of your friends: Use alcohol | Yes |  |  | X |
| How many of your friends: Get drunk | Yes |  |  | X |
| How many of your friends: Smoke nicotine | Yes |  |  | X |
| How many of your friends: Use inhalants | Yes |  |  | X |
| How many of your friends: Use other substances (e.g., cocaine) | Yes |  |  | X |
| How many of your friends: Use other tobacco products like e-cigarettes, pipes, or hookah | Yes |  |  | X |
| How many of your friends: Sell drugs | Yes |  |  | X |
| How many of your friends: Have substance use problems | Yes |  |  | X |
| Ever had a positive hair toxicology test | No |  |  | X |
| Visit type (in-person, remote, hybrid) | Yes |  |  | X |

Sensitivity weighting models were run to assess potential for noise. First, we assessed whether running mutually exclusive v. inclusive models (e.g., including sociodemographics in each model v. including other weights) created a more coherent modeling structure. As can be seen in **Table S3**, including overlapping factors resulted in a curvilinear estimation, which does not fit expectations based on raw toxicology findings, self-reported use, or outside epidemiological data. We therefore chose to use largely mutually exclusive factors in each weight.

**Table S3**. THCCOOH Estimation when including overlapping variables in each individual weight

| **THCCOOH Positive** | **High Threshold Weighted**  **% (CI)** |
| --- | --- |
| Baseline (n=432) | 3.3% (1.5-12.0) |
| Wave 1 (n=663) | 2.8% (1.6-5.1) |
| Wave 2 (n=3,335) | 3.0% (1.4-6.5) |
| Wave 3 (n=317) | 5.8% (2.4-13.4) |
| Wave 4 (n=2,803) | 6.4% (5.3-7.7) |
| Wave 5 (n=2,087) | 5.0% (4.1-6.2) |
| Wave 6 (n=1,970) | 4.7% (4.0-5.5) |

We also ran sensitivity analyses to assess whether or not trimming individual rates increased or decreased noise in estimation models. In **Table S4**, we trim (at the 95^th^ percentile) the Wave Participation Weight and Hair Weight prior to calculating the product with Baseline Weight.

**Table S4.** THCCOOH Estimation without overlapping factors and including trimmed wave participation and hair weight.

| **THCCOOH Positive** | **High Threshold Weighted**  **% (CI)** |
| --- | --- |
| Baseline (n=432) | 2.2% (0.7-6.9) |
| Wave 1 (n=663) | 3.2% (1.8-5.7) |
| Wave 2 (n=3,335) | 2.1% (1.6-2.8) |
| Wave 3 (n=317) | 4.5% (2.5-8.1) |
| Wave 4 (n=2,803) | 6.2% (5.3-7.4) |
| Wave 5 (n=2,087) | 5.9% (4.8-7.2) |
| Wave 6 (n=1,970) | 6.8% (5.8-8.0) |

In **Table S5**, we trim (at the 95^th^ percentile) the Wave Participation Weight and Hair Weight prior to calculating the product with Baseline Weight, then trim the product as well.

**Table S5.** THCCOOH Estimation without overlapping factors and including trimmed wave participation and hair weight, and trim the product of wave*hair*Baseline.

| **THCCOOH Positive** | **High Threshold Weighted**  **% (CI)** |
| --- | --- |
| Baseline (n=432) | 2.2% (0.7-6.9) |
| Wave 1 (n=663) | 3.3% (1.8-5.8) |
| Wave 2 (n=3,335) | 2.2% (1.7-2.9) |
| Wave 3 (n=317) | 4.5% (2.5-8.1) |
| Wave 4 (n=2,803) | 6.4% (5.4-7.6) |
| Wave 5 (n=2,087) | 5.9% (4.9-7.2) |
| Wave 6 (n=1,970) | 6.9% (5.9-8.1) |

To assess range of weight values, the final weights used in prevalence estimates were displayed as boxplots by wave (continued onto the next page).

**Figure S1. Boxplots of Weight Values by Wave**

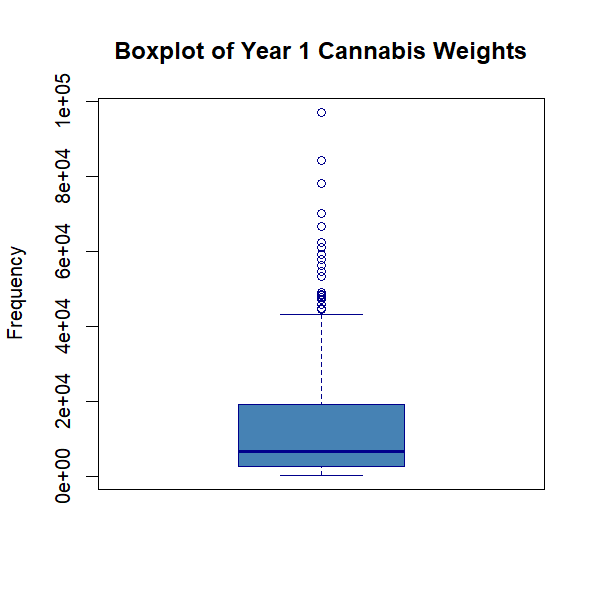

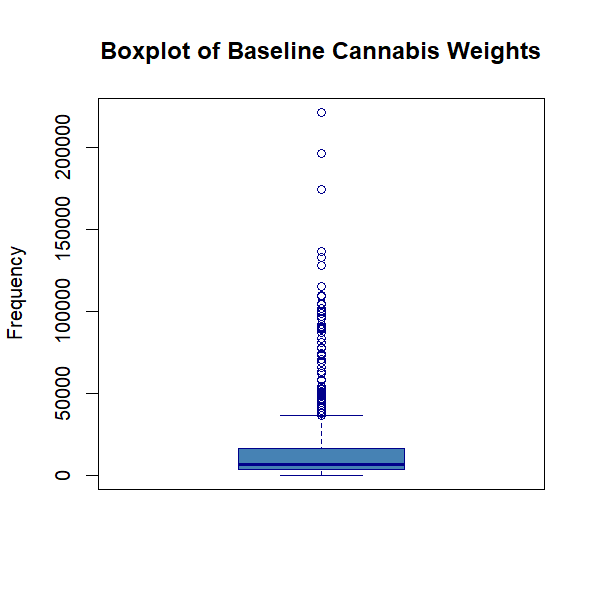

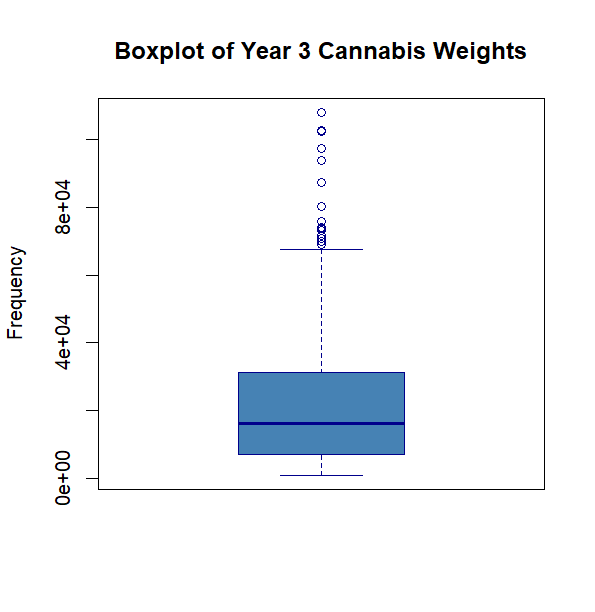

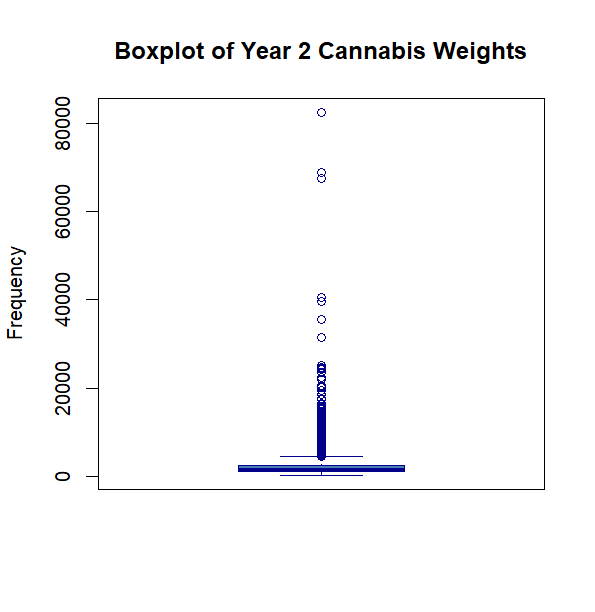

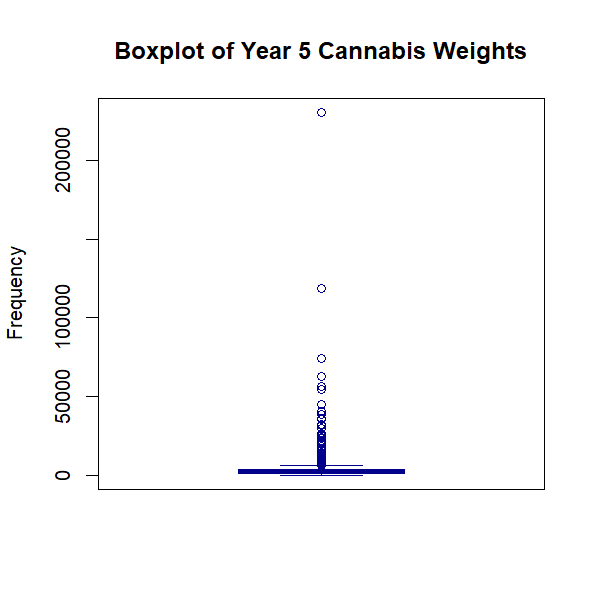

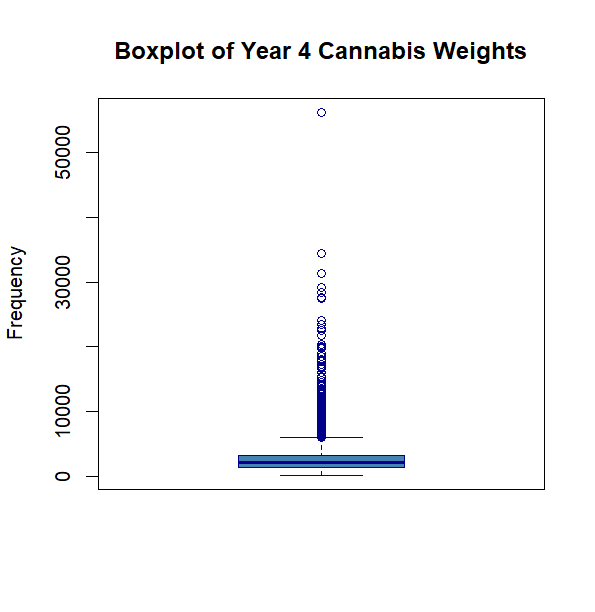

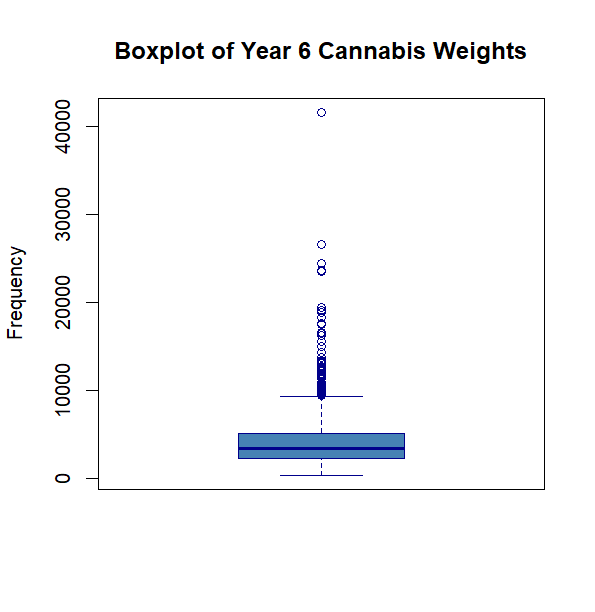

**Cannabinoid Positive Hair Tests**

Data here is for any cannabinoid positives (i.e., THCCOOH, THC, or CBD), indicating any cannabis exposure. THCCOOH positives in hair are viewed as definitive evidence of personal use of cannabinoid products, offering an even more conservative estimate of cannabis exposure than inclusion of parent analytes (e.g., THC, CBD) which can be present in cannabis smoke. **Table S6** displays estimated prevalence of any cannabinoid positives across waves.

**Table S6**. Weighted Prevalence Estimates of Cannabinoids.

|  | **High Threshold Unweighted % (CI)** | **High Threshold Weighted**  **% (CI)** | **Low Threshold**  **Unweighted**  **% (CI)** | | **Low Threshold**  **Weighted**  **% (CI)** |
| --- | --- | --- | --- | --- | --- |
| **Cannabinoid Positive** | | | |  | |
| Baseline (n=432) | 3.5% (1.8-5.2) | 2.6% (1.0-7.1) | 5.3% (3.2-7.4) | | 4.7% (2.1-10.2) |
| Wave 1 (n=663) | 3.6% (2.2-5.0) | 3.6% (2.1-6.2) | 8.9% (6.7-11.1) | | 8.1% (5.7-11.4) |
| Wave 2 (n=3,335) | 2.9% (2.3-3.5) | 2.3% (1.8-3.0) | 5.3% (4.5-6.0) | | 4.2% (3.5-5.2) |
| Wave 3 (n=317) | 7.6% (4.7-10.5) | 5.0% (2.8-8.6) | 16.4% (12.3-20.5) | | 9.9% (6.8-14.4) |
| Wave 4 (n=2,803) | 8.7% (7.7-9.8) | 7.1% (6.0-8.3) | 12.1% (10.9-13.3) | | 9.6% (8.3-11.1) |
| Wave 5 (n=2,087) | 11.8% (10.4-13.2) | 6.5% (5.3-8.0) | 13.2% (11.8-14.7) | | 7.5% (6.2-9.1) |
| Wave 6 (n=1,970) | 11.4% (10-12.8) | 7.3% (6.2-8.6) | 11.6% (10.2-13.0) | | 7.4% (6.3-8.7) |

**Weighted Prevalence of Wave Participation*Hair Test without Baseline Weight**

In order to assess generalizability of hair samples to the full ABCD cohort, rather than estimating national prevalence, a final weight was re-run and calculated for the product of Wave Participation * Hair Test. The wave participation weight included relevant sociodemographic variables which had previously been included only through the Baseline weights in the logistic regression model. As can be seen in Table S7 and relative to Figure 2 or Table S11, estimated prevalence follows the same general pattern, albeit slightly attenuated and with a higher confidence interval for some waves (e.g., Wave 3).

**Table S7.** THCCOOH Estimation Without Baseline Weight.

| **THCCOOH Positive** | **High Threshold Weighted**  **% (CI)** |
| --- | --- |
| **Baseline (n=432)** | 1.7% (0.6-4.7) |
| **Wave 1 (n=663)** | 2.4% (1.4-4.5) |
| **Wave 2 (n=3,335)** | 2.1% (1.6-2.9) |
| **Wave 3 (n=317)** | 7.1% (3.4-14.3) |
| **Wave 4 (n=2,803)** | 5.4% (4.6-6.3) |
| **Wave 5 (n=2,087)** | 5.3% (4.4-6.5) |
| **Wave 6 (n=1,970)** | 6.8% (5.7-8.0) |

**Number of samples tested and insufficient samples**

The total number of samples, by laboratory, can be seen in **Table S8**. Beyond the total number of samples sent for testing, there were insufficient quantities of hair remaining to complete all requested tests (either screening or confirmation). These numbers varied by drug class. For cannabis, there were n=258 samples across waves that had insufficient hair quantity to undergo screening or confirmation testing, and 216 of those were presumptive positives on screening which could not complete confirmation testing. For alcohol, n=551 had insufficient quantity for testing. For nicotine, n=145 presumptive positives could not undergo confirmation testing.

**Table S8. Number of Samples Tested by Wave by Lab**

|  | **All samples**  **(% of participants)** | **Psychemedics**  **(% of tested samples)** | **USDTL**  **(% of tested samples)** |
| --- | --- | --- | --- |
|  | N (%) | N (%) | N (%) |
| **Baseline (Y0)** | 438 (3.7%) | 433 (98.8%) | <10 (<2%) |
| **Wave 1 (Y1)** | 686 (6.1%) | 680 (99.1%) | <10 (<1%) |
| **Wave 2 (Y2)** | 3,394 (30.9%) | 1,102 (32.5%) | 2,292 (67.5%) |
| **Wave 3 (Y3)** | 326 (3.1%) | 281 (86.2%) | 45 (13.8%) |
| **Wave 4 (Y4)** | 2,900 (29.8%) | 1,007 (34.7%) | 1,893 (65.3%) |
| **Wave 5 (Y5)** | 2,127 (23.9%) | 413 (19.4%) | 1,714 (80.6%) |
| **Wave 6 (Y6)** | 1,996 (39.5%) | 20 (0.01%) | 1,976 (99.9%) |

**All Drug Class Positives**

Though not explored in detail here, all hair samples underwent a full drug panel for screening and, on presumptive positive, confirmation testing. **Table S9** details the number of positives by drug class as determined by each lab using their own lab cutoffs.

**Table S9.**

| **Drug Class** | **N** | **%** |
| --- | --- | --- |
| **Cannabis** | 868 | 7.3% |
| **Nicotine** | 303 | 2.6% |
| **Cocaine** | 146 | 1.2% |
| **Alcohol** | 124 | 1.0% |
| **Amp/Meth** | 85 | 0.7% |
| **Fentanyl** | 12 | 0.1% |
| **Opiates** | <10 | 0.1% |
| **Benzos** | <10 | 0.0% |
| **PCP** | 0 | 0.0% |
| **Any Drug** | 1,278 | 10.8% |

**Table S10. Sociodemographics by Wave of Hair Tested**

|  | **Category** | **Baseline**  **N=438** | **Wave 1**  **N=686** | **Wave 2**  **N=3393** | **Wave 3**  **N=326** | **Wave 4**  **N=2900** | **Wave 5**  **N=2126** | **Wave 6**  **N=1996** |
| --- | --- | --- | --- | --- | --- | --- | --- | --- |
| **Sex** | | | | | | | | |
|  | M | 53.6% | 53.5% | 46.6% | 47.9% | 44.7% | 44.6% | 46.4% |
|  | F | 46.3% | 46.5% | 53.4% | 52.1% | 55.3% | 55.4% | 53.6% |
| **Race** | | | | | | | | |
|  | White | 65.3% | 63.4% | 66.5% | 60.4% | 66.4% | 67.8% | 69.7% |
|  | Black | 6.8% | 5.4% | 4.3% | <3% | 4.0% | 3.5% | 2.8% |
|  | Asian | <1% | 1% | 1.6% | <2% | 1.6% | 1.5% | 1.2% |
|  | Hispanic | 17.1% | 18.7% | 17.6% | 23.0% | 17.3% | 16.5% | 17.6% |
|  | Other | 10.3% | 11.5% | 10.0% | 12.3% | 10.7% | 10.7% | 8.7% |
| **Family Income** | | | | | | | | |
|  | <$25K | 13.7% | 10.5% | 8.1% | 12.0% | 6.8% | 5.0% | 4.5% |
|  | $25K-$49K | 16.7% | 13.0% | 12.0% | 13.8% | 9.5% | 8.0% | 9.5% |
|  | $50K-$74K | 14.6% | 11.8% | 14.6% | 16.0% | 12.0% | 10.3% | 9.2% |
|  | $75K-$99K | 13.9% | 13.8% | 14.5% | 13.5% | 14.1% | 12.2% | 13.1% |
|  | $100K-$199K | 26.7% | 34.5% | 36.0% | 28.8% | 37.8% | 41.5% | 41.0% |
|  | $200K + | 14.4% | 16.3% | 14.7% | 16.0% | 19.8% | 23.0% | 22.7% |
| **Region** | | | | | | | | |
|  | Northeast | 14.2% | 14.0% | 12.8% | 12.3% | 16.4% | 17.1% | 16.1% |
|  | Midwest | 18.9% | 21.3% | 19.5% | 17.5% | 17.4% | 20.1% | 19.9% |
|  | South | 29.2% | 24.8% | 31.0% | 25.5% | 30.6% | 29.7% | 28.5% |
|  | West | 37.7% | 39.9% | 36.7% | 44.8% | 35.6% | 33.1% | 35.4% |

**Figure S1. Confusion Matrices of Self-Report and Toxicology by Wave and Substance**

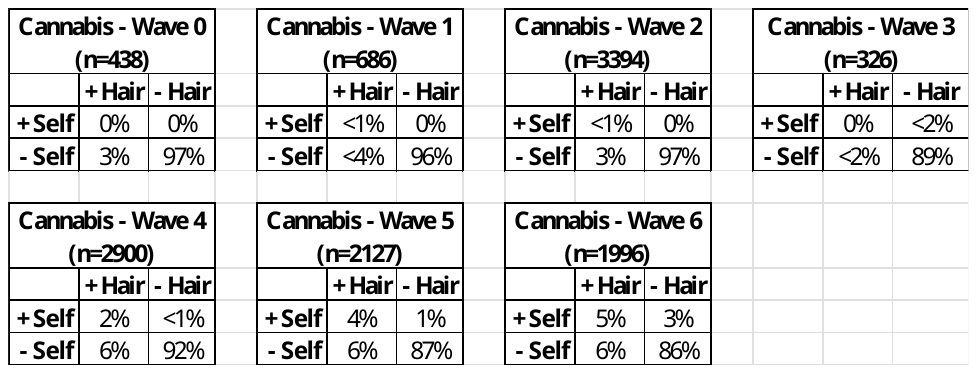

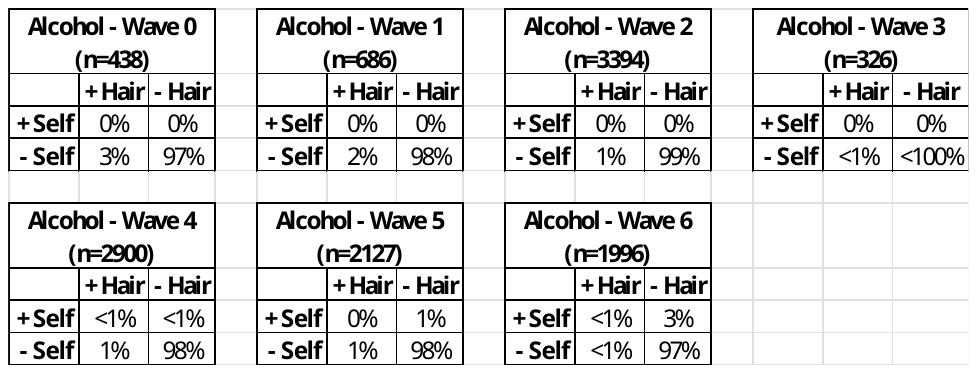

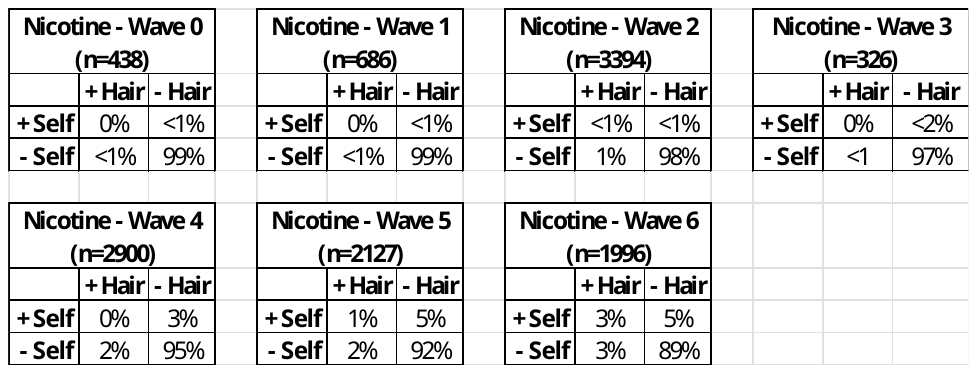

**Notes:** All concordance rates are unweighted, with raw data presented. + Hair indicates: positive hair sample based on each labs cutoff for confirmation; - Hair: negative hair sample; + Self: self-reported substance use of 6 use days in the past 3 months for cannabis and alcohol, and 3 use days in the past 3 months for nicotine; - Self: less than 6 the required use days self-reported. Only percentages are included due to some cells having small cell sizes. Consistent with recommended reporting standards from NIDA for ABCD data, n’s < 10 are obscured.

**Table S11. Concordance between Hair EtG and Self-Report by Wave (n_samples_=11,865)**

|  | **% Self-Reported Alcohol** | **% + Hair EtG** | **% Concordance** | **% Positive + Concordant/**  **EtG** |
| --- | --- | --- | --- | --- |
| Baseline | 0% | 3.4% | 96.5% | 0% |
| Wave 1 | 0% | 1.9% | 98.1% | 0% |
| Wave 2 | 0% | 0.7% | 99.4% | 0% |
| Wave 3 | 0% | <1% | 99.4% | 0% |
| Wave 4 | <1% | 1.3% | 98.4% | <10% |
| Wave 5 | 1.2% | 1.2% | 97.7% | 0% |
| Wave 6 | 2.6% | <1% | 97.1% | <20% |

Notes: All concordance rates are unweighted. % Self-Report is the percentage of participants who self-reported alcohol use ≥2x/month by wave; % + Hair EtG is the percentage of samples tested that were positive for cotinine on confirmation testing; % Concordant/All Samples represent percentage of youth whose hair results matched their self-report (either positive hair results and self-reported, or negative hair results and denied use); % Positive + Concordant/Positive Cotinine shows that same percentage, but is restricted to those who are concordant who tested positive for EtG. The lab-determined positive hair testing identification was used here. Consistent with recommended reporting standards from NIDA for ABCD data, n’s < 10 are obscured.

**Table S12**. Concordance between Hair Cotinine and Self-Report by Wave (n_samples_=11,865)

|  | **% Self-Reported Nicotine** | **% + Hair Cotinine** | **% Concordance** | **% Positive + Concordant/**  **Cotinine** |
| --- | --- | --- | --- | --- |
| Baseline | <1% | <1% | 99.3% | 0% |
| Wave 1 | <1% | <1% | 99.4% | 0% |
| Wave 2 | <2% | <1% | 98.4% | <1% |
| Wave 3 | <5% | <1% | 97.2% | 0% |
| Wave 4 | 3% | 1.9% | 95.8% | 18.5% |
| Wave 5 | 5.6% | 3.1% | 93.1% | 42.6% |
| Wave 6 | 8.2% | 6.2% | 91.4% | 46.3% |

Notes: All concordance rates are unweighted. % Self-Report is the percentage of participants who self-reported cannabis use ≥2x/month by wave; % + Hair Cotinine is the percentage of samples tested that were positive for cotinine on confirmation testing; % Concordant/All Samples represent percentage of youth whose hair results matched their self-report (either positive hair results and self-reported, or negative hair results and denied use); % Positive + Concordant/Positive Cotinine shows that same percentage, but is restricted to those who are concordant who tested positive for cotinine. The lab-determined positive hair testing identification was used here. Consistent with recommended reporting standards from NIDA for ABCD data, n’s < 10 are obscured.

**Full Weighted Prevalence Results by Substance and Wave**

Full weighted estimates are provided in Table S13, including both high and low thresholds for detection of drug analytes in hair, with a figure display of results for low threshold estimates in Figure S1. While the thresholds are actually the same for THCCOOH, there is a larger range between high and low threshold results for alcohol and nicotine. This range is due to the higher sensitivity of the low threshold results, balanced by the higher specificity of the higher threshold results. Actual prevalence may likely fall between these two ranges.

**Table S13. Full Weighted Prevalence Results by Substance and Wave**

|  | **High Threshold Unweighted % (CI)** | | **High Threshold Weighted**  **% (CI)** | | **Low Threshold**  **Unweighted**  **% (CI)** | | **Low Threshold**  **Weighted**  **% (CI)** |
| --- | --- | --- | --- | --- | --- | --- | --- |
| **THCCOOH Positive** | | | | | |  | |
| Baseline (n=432) | 3.0% (1.4-4.6) | | 2.2% (0.7-6.9) | | 3.0% (1.4-4.6) | | 2.2% (0.7-6.9) |
| Wave 1 (n=663) | 3.2% (1.8-4.5) | | 3.3% (1.8-5.8) | | 3.2% (1.8-4.5) | | 3.3% (1.8-5.8) |
| Wave 2 (n=3,335) | 2.5% (2.0-3.0) | | 2.1% (1.6-2.8) | | 2.5% (2.0-3.0) | | 2.1% (1.6-2.8) |
| Wave 3 (n=317) | 6.0% (3.4-8.6) | | 4.6% (2.5-8.2) | | 6.0% (3.4-8.6) | | 4.6% (2.5-8.2) |
| Wave 4 (n=2,803) | 7.9% (6.9-8.9) | | 6.4% (5.3-7.6) | | 7.9% (6.9-8.9) | | 6.4% (5.3-7.6) |
| Wave 5 (n=2,087) | 10.8% (9.5-12.2) | | 6.0% (4.9-7.4) | | 10.9% (9.5-12.2) | | 6.0% (4.9-7.4) |
| Wave 6 (n=1,970) | 11.2% (9.8-12.6) | | 7.1% (6.0-8.3) | | 11.2% (9.8-12.6) | | 7.1% (6.0-8.3) |
| **Alcohol Positive** | |  | |  | |  | |
| Baseline (n=438) | 0.2% (0.0-0.7) | | 0.3% (0.0-2.3) | | 5.3% (3.2-7.3) | | 3.5% (1.4-8.3) |
| Wave 1 (n=686) | 0.2% (0.0-0.4) | | 0.1% (0.0-0.1) | | 3.2% (1.9-4.5) | | 3.0% (1.7-5.2) |
| Wave 2 (n=3,302) | 0.2% (0.1-0.4) | | 0.1% (0.0-0.3) | | 1.3% (0.9-1.6) | | 0.9% (0.6-1.3) |
| Wave 3 (n=322) | 0.3% (0.0-0.9) | | 1.0% (0.1-6.7) | | 1.6% (0.2-2.9) | | 2.0% (0.7-5.9) |
| Wave 4 (2,784) | 0.4% (0.2-0.6) | | 0.2% (0.1-0.4) | | 2.0 % (1.5-2.5) | | 1.3% (0.9-1.7) |
| Wave 5 (n=1,996) | 0.8% (0.4-1.1) | | 0.4% (0.2-0.7) | | 1.6% (1.1-2.2) | | 0.8% (0.5-1.3) |
| Wave 6 (n=1,845) | 0.3% (0.1-0.6) | | 0.2% (0.1-0.4) | | 0.5% (0.2-0.8) | | 0.3% (0.1-0.6) |
| **Nicotine Positive** | |  | |  | |  | |
| Baseline (n=436) | 0.0% (0.0-0.0) | | 0.0% (0.0-0.0) | | 0.2% (0.0-0.7) | | 0.03% (0.0-0.2) |
| Wave 1 (n=683) | 0.2% (0.0-0.4) | | 0.1 % (0.0-0.9) | | 0.4% (0.0-0.9) | | 0.5% (0.1-1.5) |
| Wave 2 (n=3,370) | 1.4% (1.0-1.8) | | 1.1% (0.7-1.6) | | 1.6% (1.2-2.0) | | 1.2% (0.9-1.8) |
| Wave 3 (n=325) | 0.6% (0.0-1.5) | | 0.3% (0.1-1.1) | | 0.6% (0.0-1.5) | | 0.3% (0.1-1.1) |
| Wave 4 (n=2,865) | 1.5% (1.1-2.0) | | 1.4% (0.9-2.0) | | 1.8% (1.3-2.3) | | 1.6% (1.1-2.3) |
| Wave 5 (n=2,089) | 2.8% (2.1-3.5) | | 1.7% (1.1-2.5) | | 3.3% (2.5-4.0) | | 1.9% (1.3-2.8) |
| Wave 6 (n=1,952) | 6.3% (5.2-7.4) | | 4.7% (3.7-6.0) | | 6.3% (5.2-7.4) | | 4.7% (3.7-6.0) |

Notes: n’s represent the number of samples which were assayed for each drug class by wave; high threshold refers to having a higher cutoff to determine a positive hair test, resulting in a more conservative estimate; low threshold refers to having a cutoff set to the level of detection (LOD) of the lab to identify a positive hair test, resulting in a more sensitive estimate

**Figure S2. Low Threshold Weighted Prevalence Estimates by Substance and Wave**

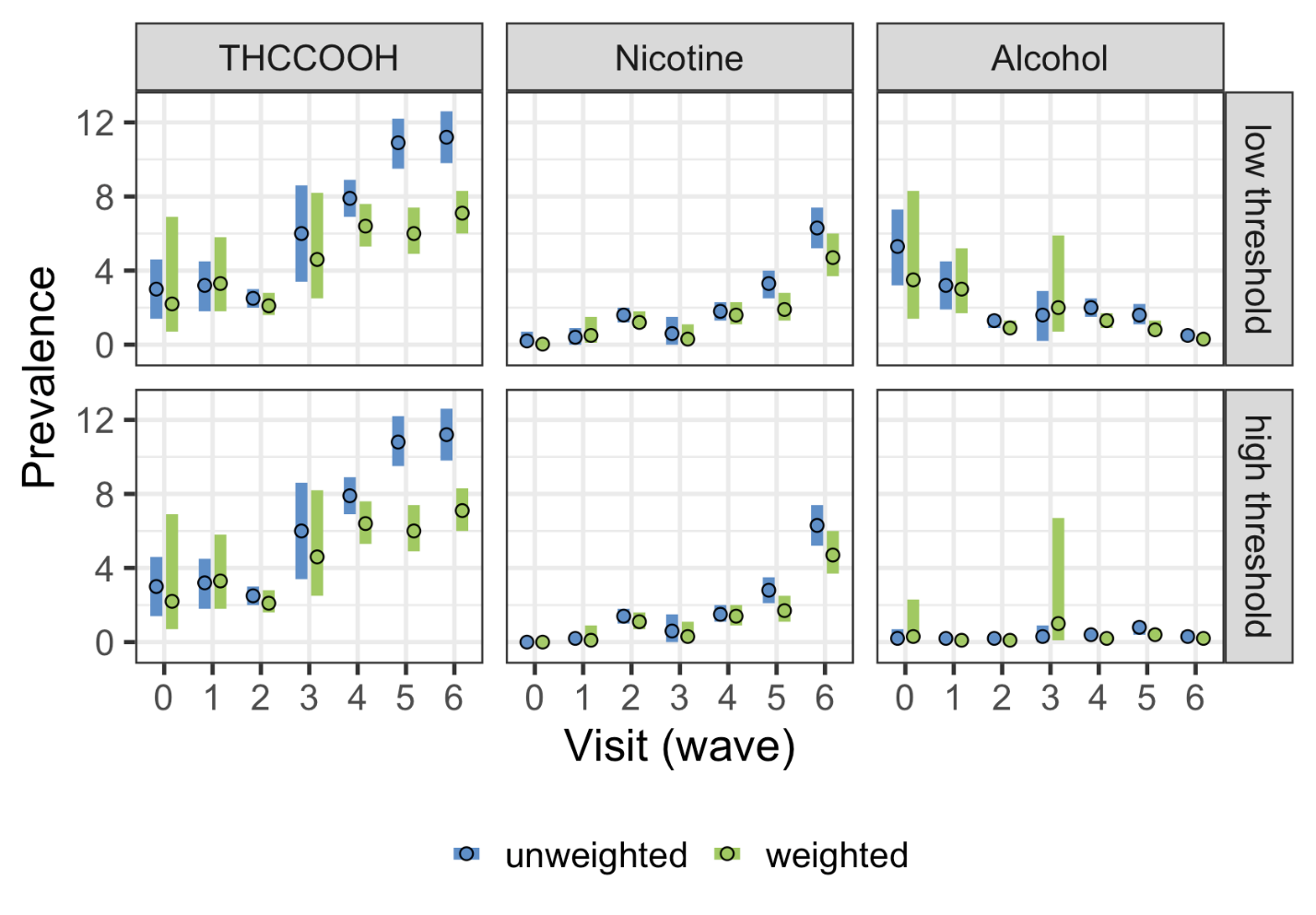

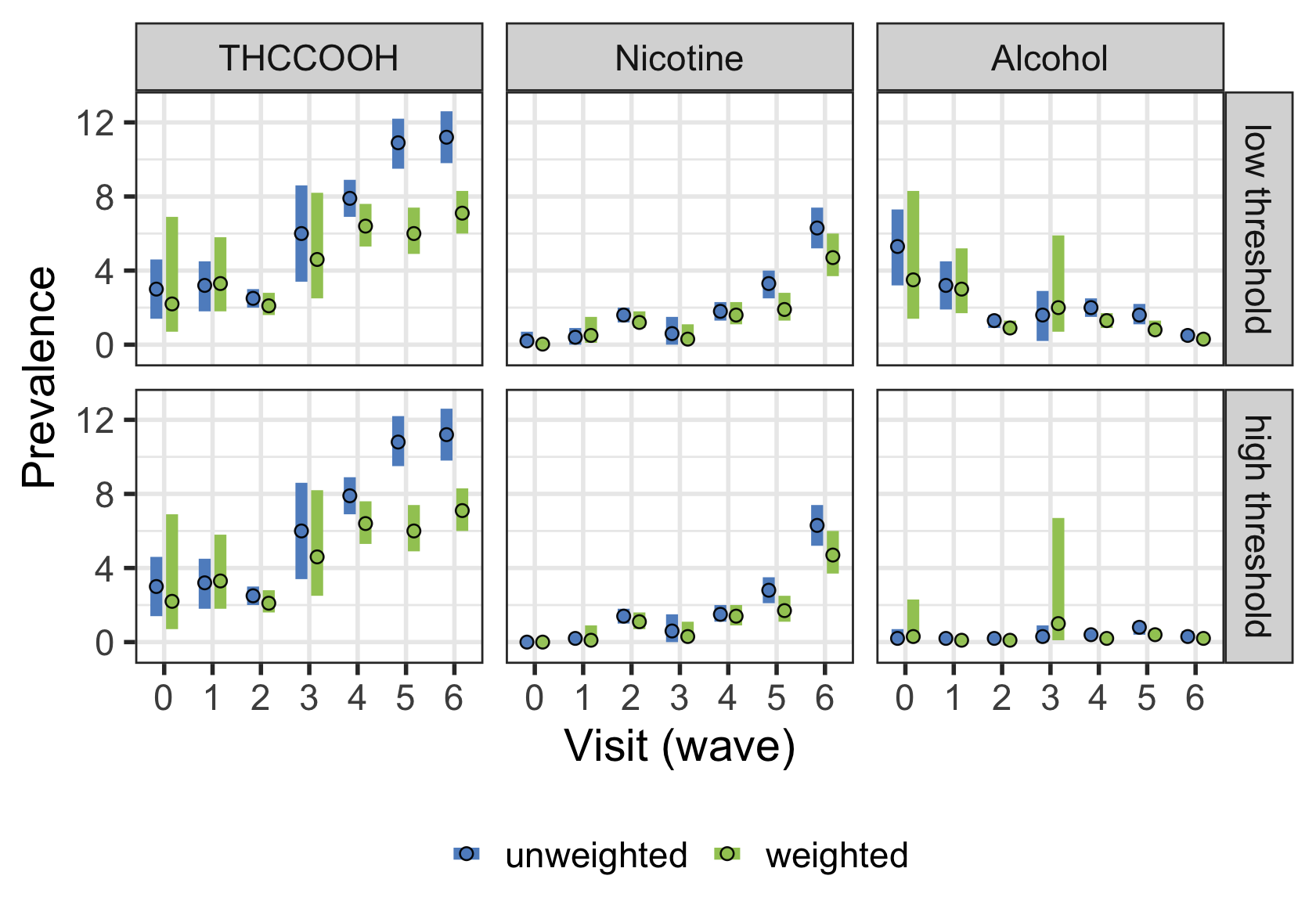

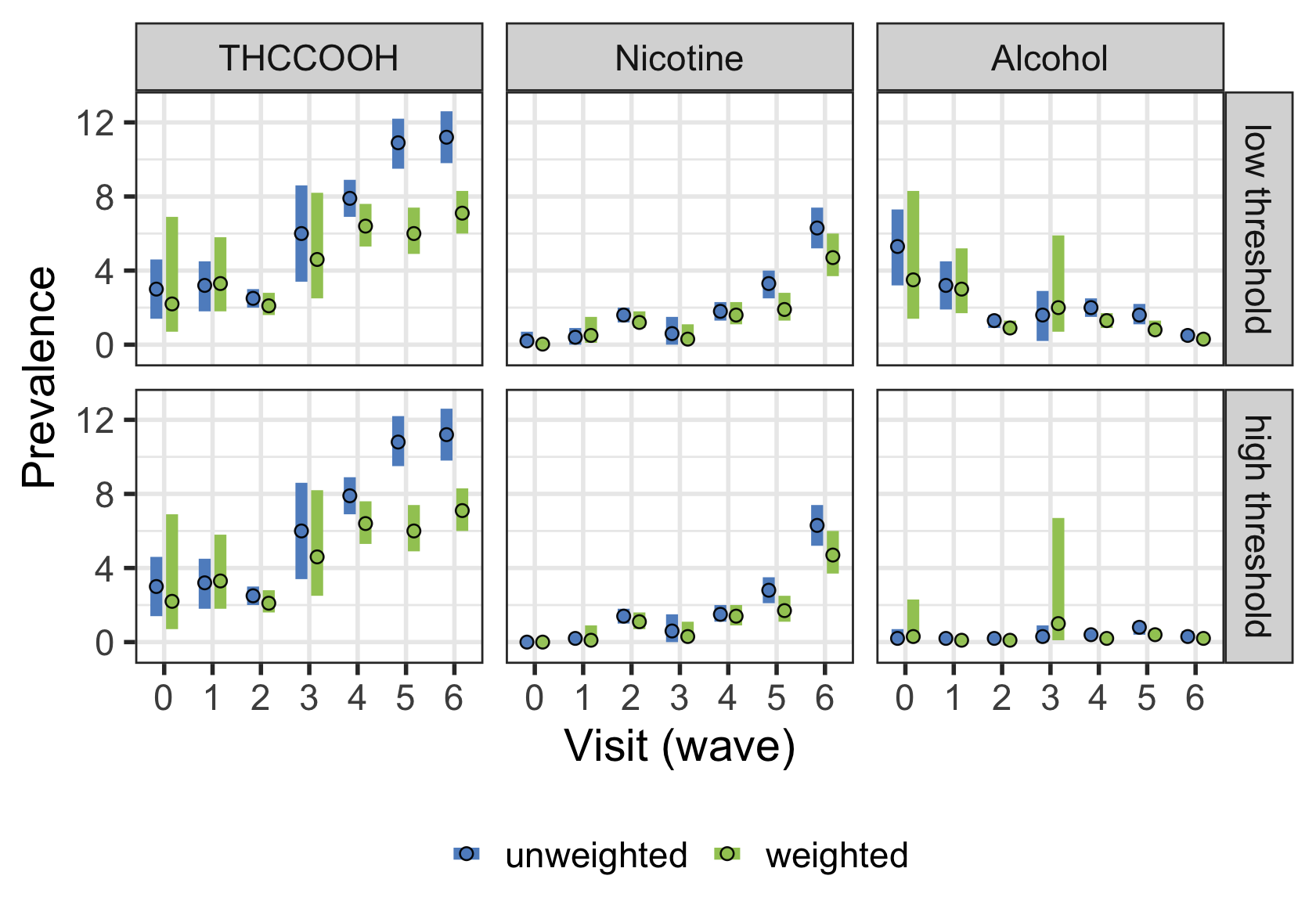

Notes: Shaded regions represent 95% confidence interval; low threshold refers to a cutoff set at the level of detection (LOD) of the lab to identify a positive hair test, resulting in a more sensitive estimate.
